# Sleep health education and a personalized smartphone application improve sleep and productivity and reduce healthcare utilization among employees: Results of a randomized clinical trial

**DOI:** 10.1101/2021.10.13.21264974

**Authors:** Rebecca Robbins, Matthew D. Weaver, Stuart F. Quan, Jason P. Sullivan, Salim Qadri, Charles A. Czeisler, Laura Barger

## Abstract

Sleep deficiency and undiagnosed or untreated sleep disorders are pervasive among employed adults, yet often ignored in the context of workplace health promotion programs among employers. Smartphone applications (app) are a promising, scalable approach to improving sleep among employees. In this randomized clinical trial, we evaluate the dayzz app, a personalized sleep training program that promotes healthy sleep and sleep disorders awareness through personalized, comprehensive sleep improvement solutions. In a sample of daytime employees affiliated with a large healthcare organization, we evaluated the dayzz app in a parallel-group, randomized, waitlist control trial. Participants were randomly assigned to either use the dayzz app throughout the study or the waitlist control condition where they would receive the dayzz app at the end of the study period. We collected data on employee sleep (e.g., sleep duration, sleep health behavioral changes); workplace outcomes (e.g., employee presenteeism, absenteeism, and performance); and healthcare utilization (e.g., mental health, ambulatory visits, and emergency room visits), throughout the study. Results show that those assigned to the experimental condition exhibited an increase in healthy sleep behaviors; an increase in sleep duration; a trend toward a more regular sleep schedule; and a significant increase in overall sleep quality. Regarding workplace outcomes, results showed that those in the experimental condition also demonstrated a trend toward less absenteeism and significantly lower presenteeism; and those in the experimental condition reported lower healthcare utilization. Results from this randomized clinical trial demonstrate that a workplace sleep wellness program can be beneficial to both the employee and employer.

## Introduction

Seventy percent of adults in the United States (US) admit routinely obtaining insufficient sleep (less than the recommended 7 to 8 hours).^1^ Additionally, sleep disorders are very common, with 30-40% of employees screening positive for at least one common sleep disorder.^2–5^ Moreover, approximately 85% of those at risk for a sleep disorder are undiagnosed and untreated.^2,6^ Approximately 50-70 million adults in the US are at risk for a sleep disorder.^7^ Sleep deficiency and/or untreated sleep disorders are associated with significant health consequences for the individual employee, their employer, and society. Despite the importance of sufficient sleep and overall sleep health for workplace productivity as well as employee health and safety, nationally representative data collected among US employers shows that fewer than 10% of employers report that they provide sleep-focused programs for their employees, yet nearly one third report nutrition or exercise programming for employees.^8^

Previous research has shown that workplace health programs are effective for improving employee sleep, overall health, and workplace outcomes. In a randomized controlled trial of an in-person sleep education and sleep disorders screening intervention, researchers found there was a significant reduction in injuries and disability day usage.^6^ Further, among employees at a national trucking company, diagnosing and treating the sleep disorder sleep apnea through an employer-sponsored program saved the employer more than $3,000 per employee annually.^9^

Technologies, such smartphone applications (apps) hold promise for delivering sleep health and wellness information to employees and in turn, offering tangible benefits to employers in terms of improved productivity. ^10,11^ Here we report results of a randomized controlled trial evaluating sleep health education followed by a smartphone app (‘dayzz’) designed to improve sleep health among employees in workplace settings. We administered the sleep health education then provided the dayzz app to employees. We evaluated their sleep (e.g., sleep duration, sleep health behavioral changes); workplace outcomes (e.g., employee presenteeism, absenteeism, and performance); and healthcare utilization (e.g., mental health, ambulatory visits, and emergency room visits), throughout the study as compared to a waitlist control condition.

## Methods

As described in detail elsewhere,^12^ we conducted a randomized, parallel group controlled trial using a waitlist control design among daytime employees at a large healthcare organization in the US Northeast. Participants were enrolled using adaptive randomization to either experimental or control. Specifically, due to the additional steps (e.g., downloading the app) required in the experimental condition, the likelihood of being assigned to the experimental group was greater than 50% to ensure a sufficient number of participants in each group.

The experimental group received the intervention, which was comprised of: 1) sleep health education at enrollment, including screening for common sleep disorders, with those that screened positive for a sleep disorder receiving resources to enable further evaluation and treatment as appropriate over the study duration; and 2) access to the dayzz app for the study duration. The control group participants were offered the intervention at the end of the study. The study recruitment was originally intended to be conducted in-person. However, due to the onset of the COVID-19 pandemic, the study recruitment, enrollment, and follow-up procedures were re-designed to be completed entirely online.

Eligible participants were daytime workers employed by the large healthcare-oriented research and teaching organization in the US Northeast. Eligibility required participants be employed by the organization and report regularly using smartphone apps (i.e., once per week). Ineligible participants were evening, night, or rotating shift workers and those who reported being pregnant or breast-feeding.

Participants were recruited via system-wide emails to the >40,000-member organization. An advertisement for the study was also posted on a website hosted by the organization devoted to research, with a wide circulation (>10,000 visitors monthly). Finally, the study team posted targeted social media advertisements to individuals who listed the employer as their place of work.

Interested participants were directed to an online landing page with more information about the study and the opportunity to complete an online screener that automatically screened employees for eligibility. Consent was collected via electronic signature. After providing electronic consent via the online landing page, participants were randomized. Participants were compensated for their time. This study was approved by the Partners Healthcare Institutional Review Board.

After consenting, participants randomized to the experimental group took a short online initial contact questionnaire to collect basic demographic information, watched the 20-minute sleep health education video, completed a sleep disorder screener, then received information on how to download and use the dayzz app.

The dayzz app begins with a brief onboarding and registration process, followed by the administration of a digital sleep disorders questionnaire. Based on the user’s assessment, the dayzz app offers personalized sleep tips through tailored modules to deliver evidence-based therapies for the specific sleep issue users report, for example, cognitive-behavioral skills and strategies for users at risk for insomnia. All users are provided with modules covering basic sleep hygiene principles and tools, such as bedroom environment optimization (integrating smartphone noise and light sensors), white noise audios, and written or auditory content to promote sleep health.

### Measures

All participants completed the electronic sleep diary as well as monthly and end of study questionnaires, as previously described. We evaluated changes in sleep behaviors using a checklist asking participants to select the healthy sleep changes they have made since starting the study. Specifically, participants are asked “During this study, have you changed any sleep-related behaviors to improve your sleep since participating in the study (check all that apply)?” Participants had the option to select changes they may have made, such as “Go to bed earlier,” “Keep a more consistent sleeping schedule,” and “Set an alarm to remind you of your bedtime.”

Participants were also asked to self-report their sleep duration and timing on the sleep diary. The Sleep Regularity Index (SRI) is the percentage probability of an individual being in the same state (asleep vs. awake) at any two time-points 24 h apart, averaged across the sleep diary intervals.^13^ An individual who sleeps and wakes at exactly the same times each day scores 100 (better outcome), whereas an individual who sleeps and wakes at random scores 0 (worse outcome).

Participants also reported their sleep quality using the Pittsburgh Sleep Quality Index (PSQI).^14^ The PSQI differentiates “poor” from “good” sleep by measuring seven domains: subjective sleep quality, sleep latency, sleep duration, habitual sleep efficiency, sleep disturbances, use of sleep medication, and daytime dysfunction over the last month. The participant self-rated each of these seven areas of sleep.

Absenteeism, performance, and productivity was evaluated using the World Health Organization (WHO) Health and Work Performance Questionnaire Short Form.^15^ Participants were asked the number of hours worked in a typical week. Unscheduled absences and disability day usage, presenteeism, and a ratio of workplace performance relative to ideal or possible performance was evaluated. Finally, participants were asked to report interactions with the healthcare system, such as visits to the emergency room or urgent care, or to their primary care or mental health providers.

### Statistical plan

Analyses were conducted using an intention-to-treat approach (ITT). Outcome measures were compared by assignment to the experimental and waitlist control groups. The ITT analysis included all participants randomized in the study. Baseline to follow-up comparisons were conducted using the first available datapoint (96% reported in month 1 or month 2) and the mean responses from months 7-9. The distribution of the data was examined, and transformations were performed as necessary for valid comparisons. The odds of changing sleep behaviors were tested using logistic regression models. Sleep quality was assessed using the PSQI. We examined changes in sleep duration and PSQI score from baseline (first submitted survey) to follow-up (the mean of submitted values in months 7-9) in both groups. These continuous measures were compared at baseline and follow-up using two-sample t-tests. We also estimated costs due to absenteeism and presenteeism. We compared the cumulative cost of each over the study interval using two-sample t-tests. Finally, we compared monthly utilization of the healthcare system between groups using mixed models that accounted for the dependence between repeated measures. Alpha was set at 0.05 for all comparisons. Stata version 15.1 (College Station, TX) was used to conduct the statistical analysis.

## Results

The final cohort was comprised of 794 participants assigned to the experimental condition and 561 assigned to the control condition. 1,355 individuals completed 4,911 surveys over the study interval. The number of control group surveys (n=2,455) and intervention group surveys (2,456) was similar.

Age, gender, race/ethnicity, education, income, job type, and all health conditions except anxiety disorder (experimental: 13%; control: 19%, p=.01) did not vary between experimental and control conditions. See Table 1 for full demographic details describing the study sample.

**Table 1.**
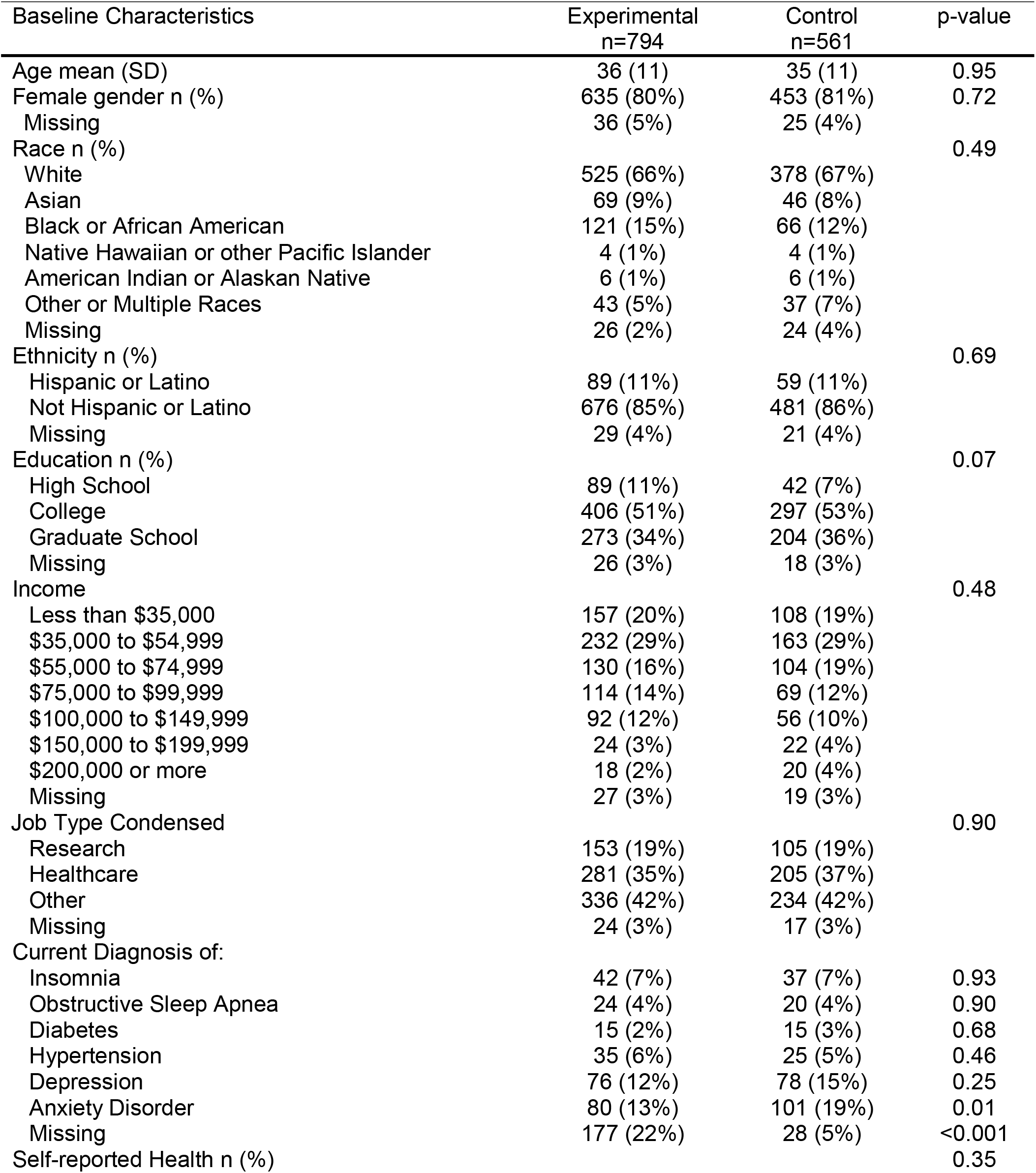
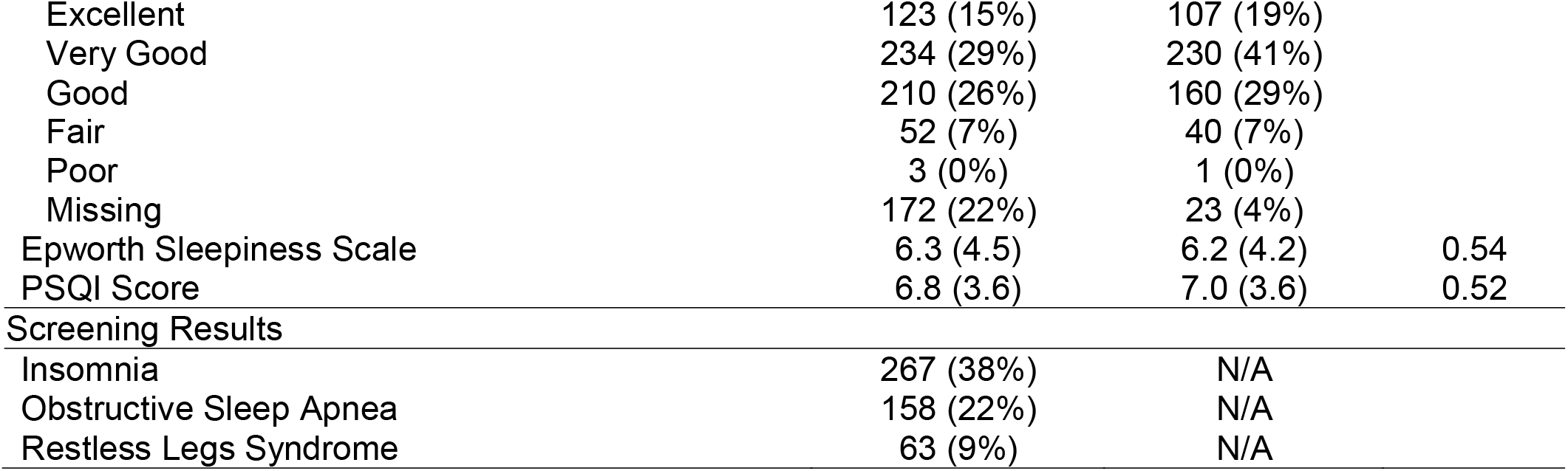
Demographic characteristics of the study sample (n=1,355).

| Baseline Characteristics | Experimental<br>n=794 | Control<br>n=561 | p-value |
| --- | --- | --- | --- |
| Age mean (SD) | 36 (11) | 35 (11) | 0.95 |
| Female gender n (%) | 635 (80%) | 453 (81%) | 0.72 |
| Missing | 36 (5%) | 25 (4%) |  |
| Race n (%) |  |  | 0.49 |
| White | 525 (66%) | 378 (67%) |  |
| Asian | 69 (9%) | 46 (8%) |  |
| Black or African American | 121 (15%) | 66 (12%) |  |
| Native Hawaiian or other Pacific Islander | 4 (1%) | 4 (1%) |  |
| American Indian or Alaskan Native | 6 (1%) | 6 (1%) |  |
| Other or Multiple Races | 43 (5%) | 37 (7%) |  |
| Missing | 26 (2%) | 24 (4%) |  |
| Ethnicity n (%) |  |  | 0.69 |
| Hispanic or Latino | 89 (11%) | 59 (11%) |  |
| Not Hispanic or Latino | 676 (85%) | 481 (86%) |  |
| Missing | 29 (4%) | 21 (4%) |  |
| Education n (%) |  |  | 0.07 |
| High School | 89 (11%) | 42 (7%) |  |
| College | 406 (51%) | 297 (53%) |  |
| Graduate School | 273 (34%) | 204 (36%) |  |
| Missing | 26 (3%) | 18 (3%) |  |
| Income |  |  | 0.48 |
| Less than \$35,000 | 157 (20%) | 108 (19%) | |
| \$35,000 to \$54,999 | 232 (29%) | 163 (29%) | |
| \$55,000 to \$74,999 | 130 (16%) | 104 (19%) | |
| \$75,000 to \$99,999 | 114 (14%) | 69 (12%) | |
| \$100,000 to \$149,999 | 92 (12%) | 56 (10%) | |
| \$150,000 to \$199,999 | 24 (3%) | 22 (4%) | |
| \$200,000 or more | 18 (2%) | 20 (4%) | |
| Missing | 27 (3%) | 19 (3%) |  |
| Job Type Condensed |  |  | 0.90 |
| Research | 153 (19%) | 105 (19%) |  |
| Healthcare | 281 (35%) | 205 (37%) |  |
| Other | 336 (42%) | 234 (42%) |  |
| Missing | 24 (3%) | 17 (3%) |  |
| Current Diagnosis of: |  |  |  |
| Insomnia | 42 (7%) | 37 (7%) | 0.93 |
| Obstructive Sleep Apnea | 24 (4%) | 20 (4%) | 0.90 |
| Diabetes | 15 (2%) | 15 (3%) | 0.68 |
| Hypertension | 35 (6%) | 25 (5%) | 0.46 |
| Depression | 76 (12%) | 78 (15%) | 0.25 |
| Anxiety Disorder | 80 (13%) | 101 (19%) | 0.01 |
| Missing | 177 (22%) | 28 (5%) | <0.001 |
| Self-reported Health n (%) |  |  | 0.35 |
| Excellent | 123 (15%) | 107 (19%) |  |
| Very Good | 234 (29%) | 230 (41%) |  |
| Good | 210 (26%) | 160 (29%) |  |
| Fair | 52 (7%) | 40 (7%) |  |
| Poor | 3 (0%) | 1 (0%) |  |
| Missing | 172 (22%) | 23 (4%) |  |
| Epworth Sleepiness Scale | 6.3 (4.5) | 6.2 (4.2) | 0.54 |
| PSQI Score | 6.8 (3.6) | 7.0 (3.6) | 0.52 |
| Screening Results |  |  |  |
| Insomnia | 267 (38%) | N/A |  |
| Obstructive Sleep Apnea | 158 (22%) | N/A |  |
| Restless Legs Syndrome | 63 (9%) | N/A |  |

### Changes in self-reported sleep behaviors

In terms of changes to sleep behaviors, 39% of participant-months reported less fatigue or sleepiness, 62% of participant-months reported increased sleep consistency, 42% reported increasing sleep duration, and 39% reported sleeping in later. The intervention group was 30% more likely to feel less fatigued or sleepy (OR 1.30; 95% CI 1.08-1.57). They were approximately 40% more likely to report increased sleep consistency (OR 1.40; 95% CI 1.12-1.75) and sleep duration (OR 1.44; 95% CI 1.17-1.78). There was no difference between groups in the odds of sleeping in later (p=0.58, Figure 1).

**Figure 1.**
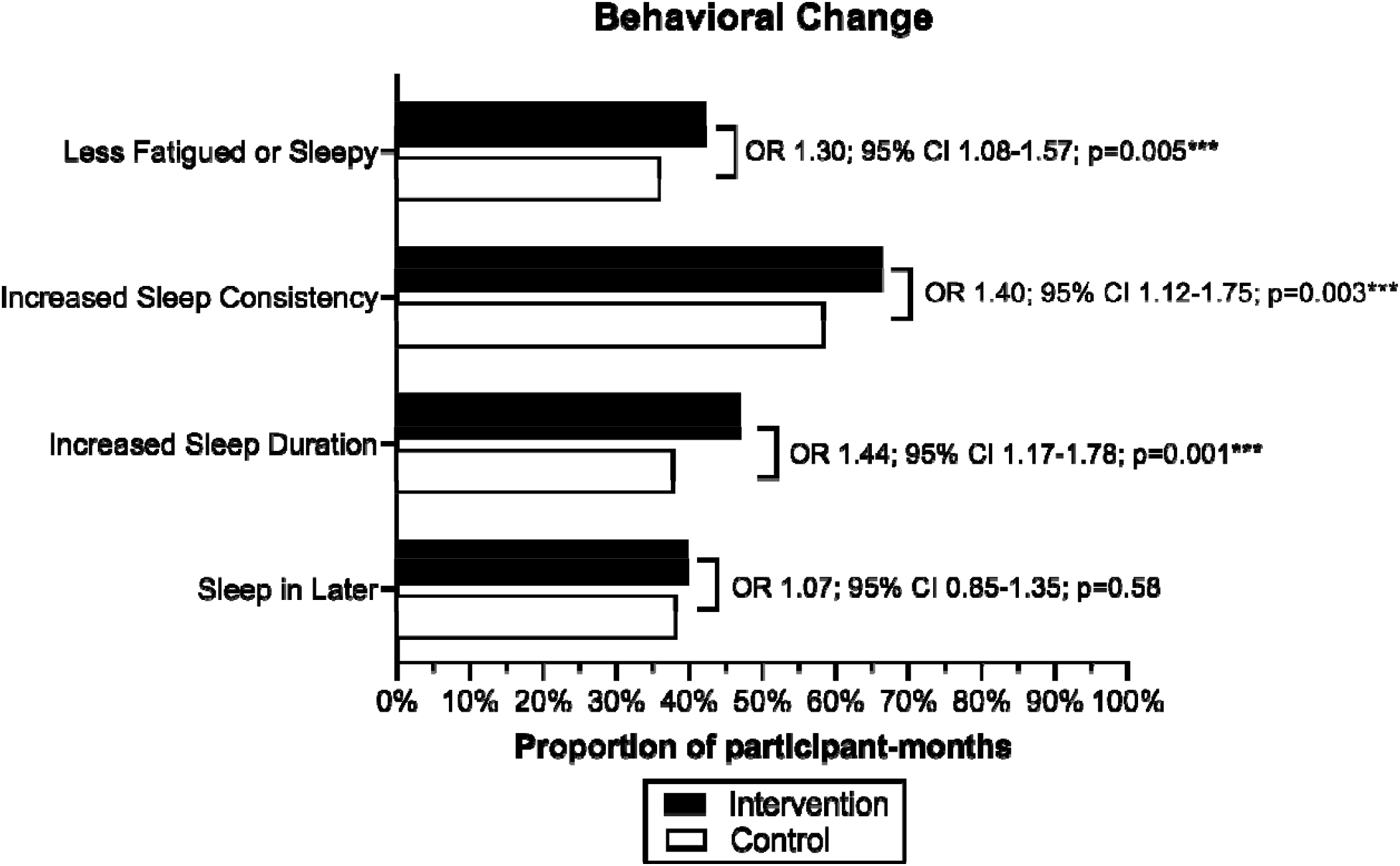
Self-reported sleep behavioral changes during the study between intervention and control conditions.

### Changes in self-reported sleep duration

The mean hours of sleep at baseline did not differ between the intervention and control groups on work days (p=0.91) or free days (p=0.57). At follow-up assessment, the intervention group reported significantly more sleep than the control group, both on work nights (p=0.01) and on free nights (p=0.03, Figure 2).

**Figure 2.**
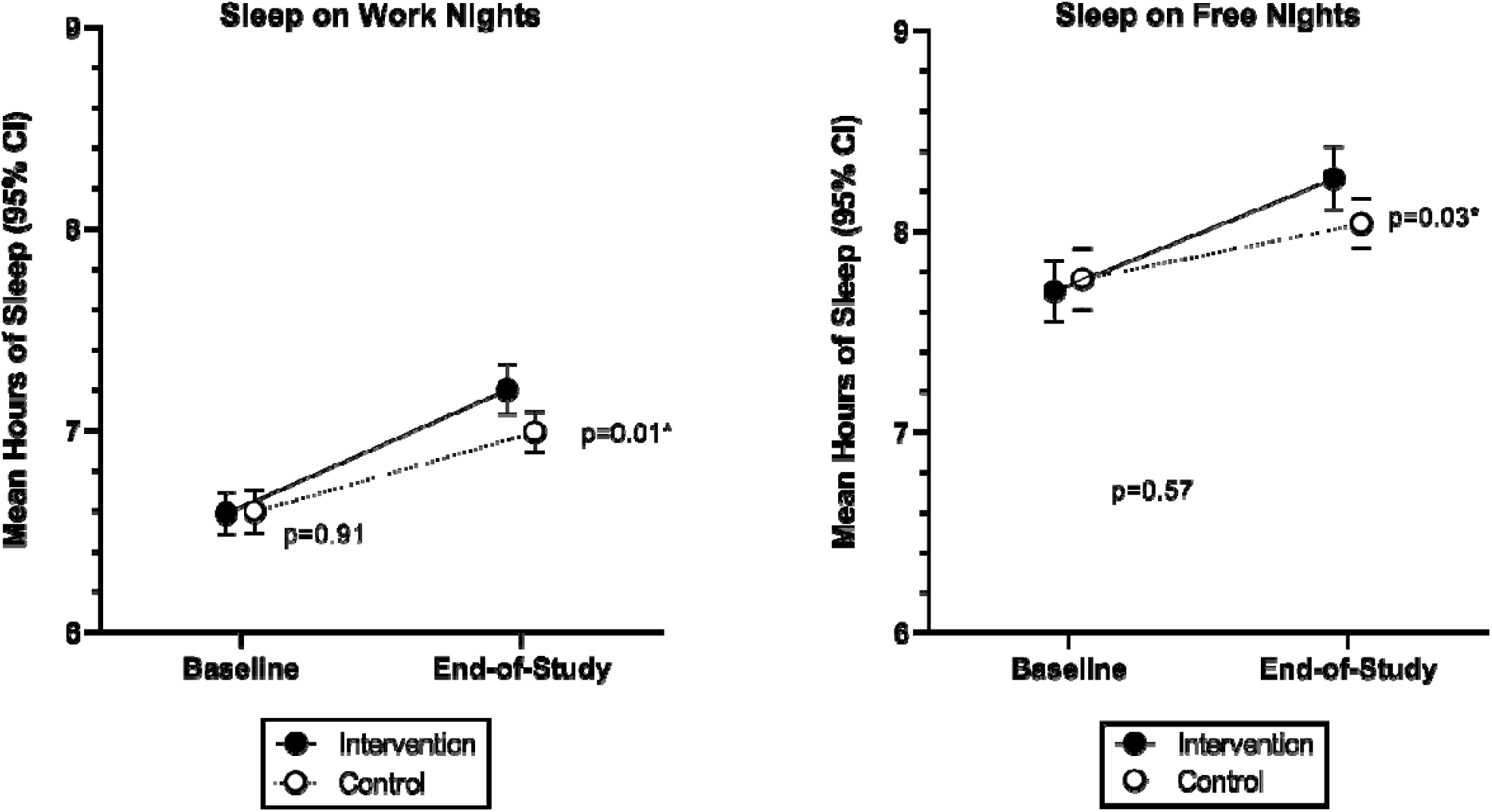
Changes to self-reported sleep duration during the study between intervention and control conditions.

### Changes in sleep quality

The mean PSQI score at baseline in the intervention group was 6.83 (95% CI 6.65-7.28), while the mean PSQI in the control group was 6.97 (95% CI 6.65-7.28). The groups did not significantly differ at baseline (p=0.52). At follow-up, the mean PSQI score in the intervention group was 5.12 (95% CI 4.88-5.36), while the mean PSQI in the control group was 5.52 (95% CI 5.29-5.76). The mean PSQI in the intervention group was significantly lower at follow-up assessment compared to the control group (p=0.02). The prevalence of poor sleep quality at baseline (PSQI >=5) was 59% in the intervention group compared to 62% in the control group (p=0.42). At follow-up, the prevalence of poor sleep quality was significantly lower in the intervention group (40%) compared to the control group (46%). The odds of poor sleep quality at follow-up was reduced by 21% in the intervention group (OR 0.79, 95% CI 0.63-0.98, Figure 3).

**Figure 3.**
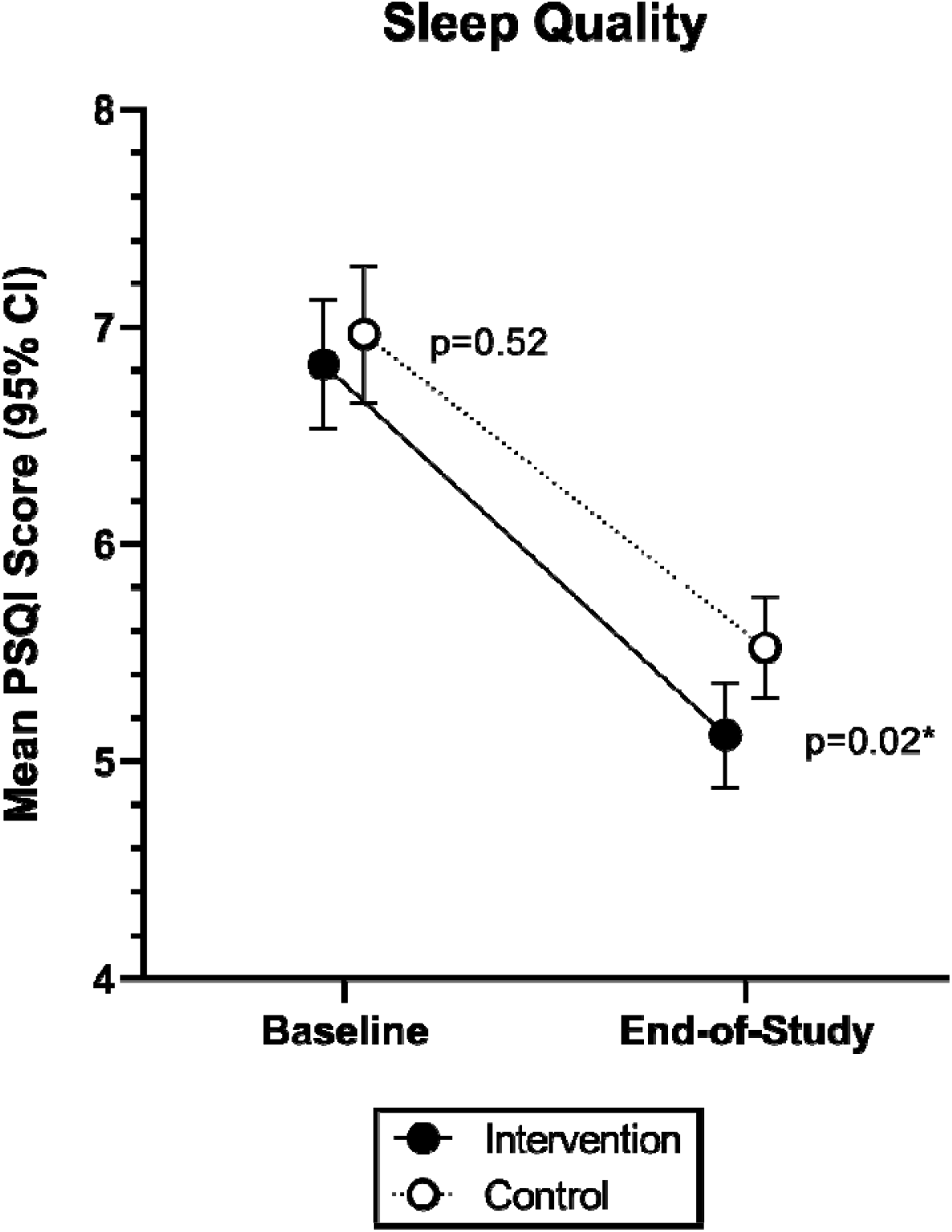
Changes to sleep quality as measured by the Pittsburgh Sleep Quality Index (PSQI) during the study between intervention and control conditions.

### Changes in presenteeism and absenteeism

The total dollars lost from absenteeism and presenteeism was calculated for each participant over the study interval. The mean total dollars lost due to absenteeism was $467 less in the experimental group, approaching statistical significance (p=0.06). The mean total dollars lost to reduced workplace performance (presenteeism) was less in the intervention group compared to the control group (p=0.0001), corresponding to an average of $2,466 dollars per person over the study interval (or $274 per month). The total lost due to presenteeism in the control group was 3.55 million dollars USD, compared to 3.07 million dollars in the experimental group, for a savings of $482,854 (Figure 4).

**Figure 4.**
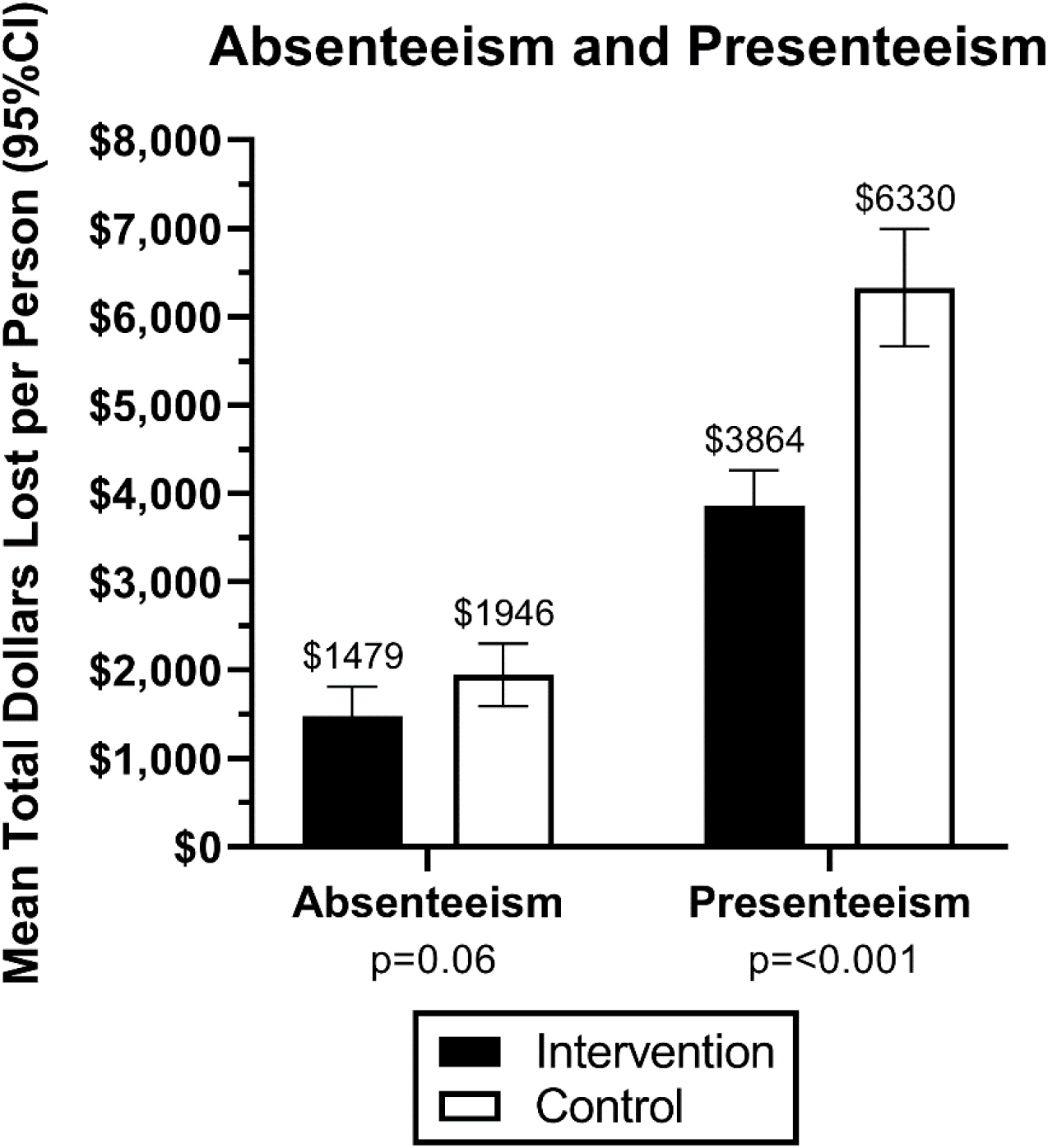
Changes in absenteeism and presenteeism during the study between intervention and control conditions.

### Changes in healthcare utilization

We observed a significant reduction in mental health visits (p=0.01) and overall healthcare utilization (p=0.03, Figure 5).

**Figure 5.**
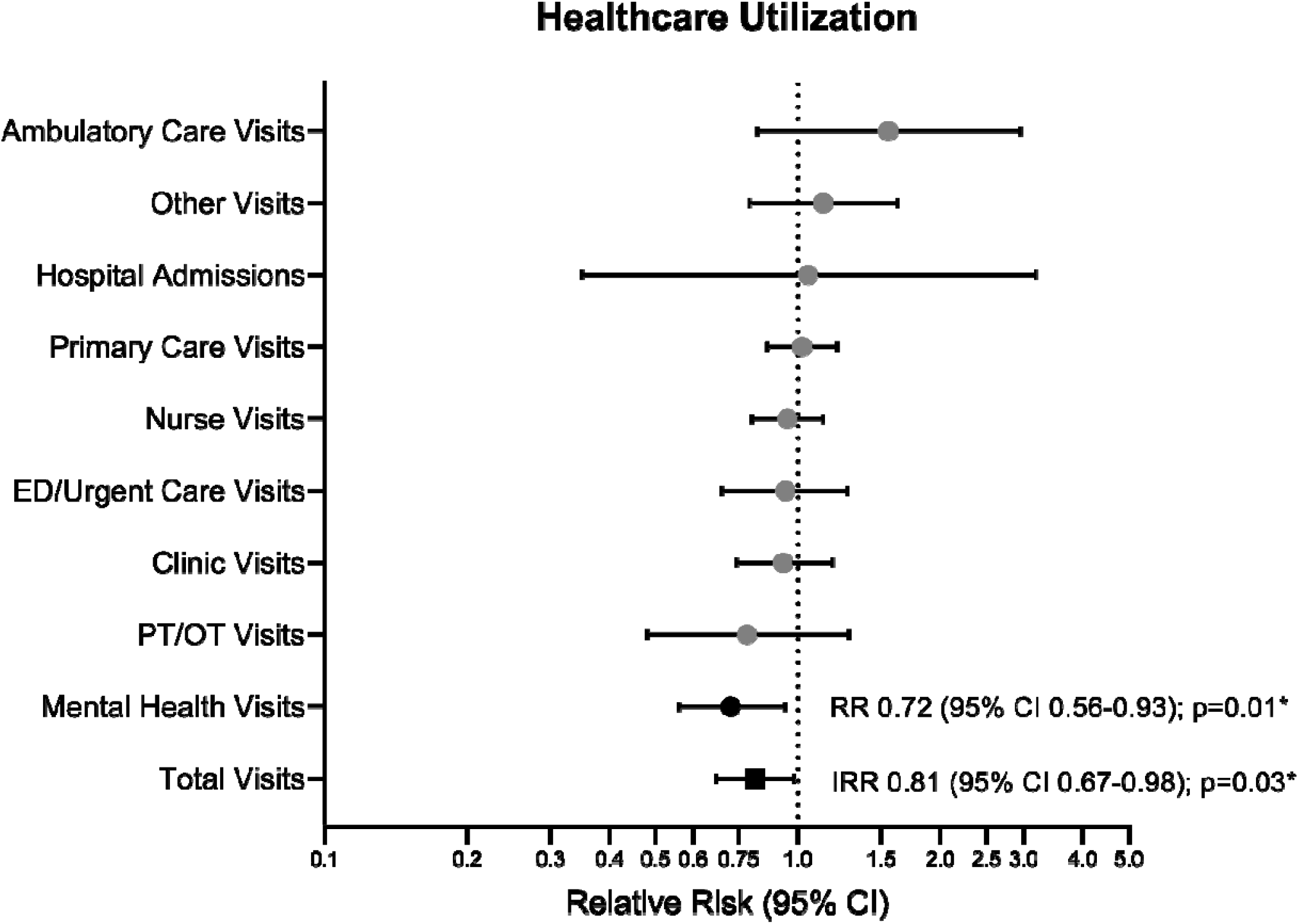
Changes in healthcare utilization during the study between intervention and control conditions.

### Changes in sleep diary-derived sleep duration

Using the sleep diary data, the Sleep Regularity Index was similar at baseline (75 in each group). On follow-up, the SRI increased to 78 in the intervention group and stayed at 75 in the control group (p=0.06 at follow-up, Figure 6).

**Figure 6.**
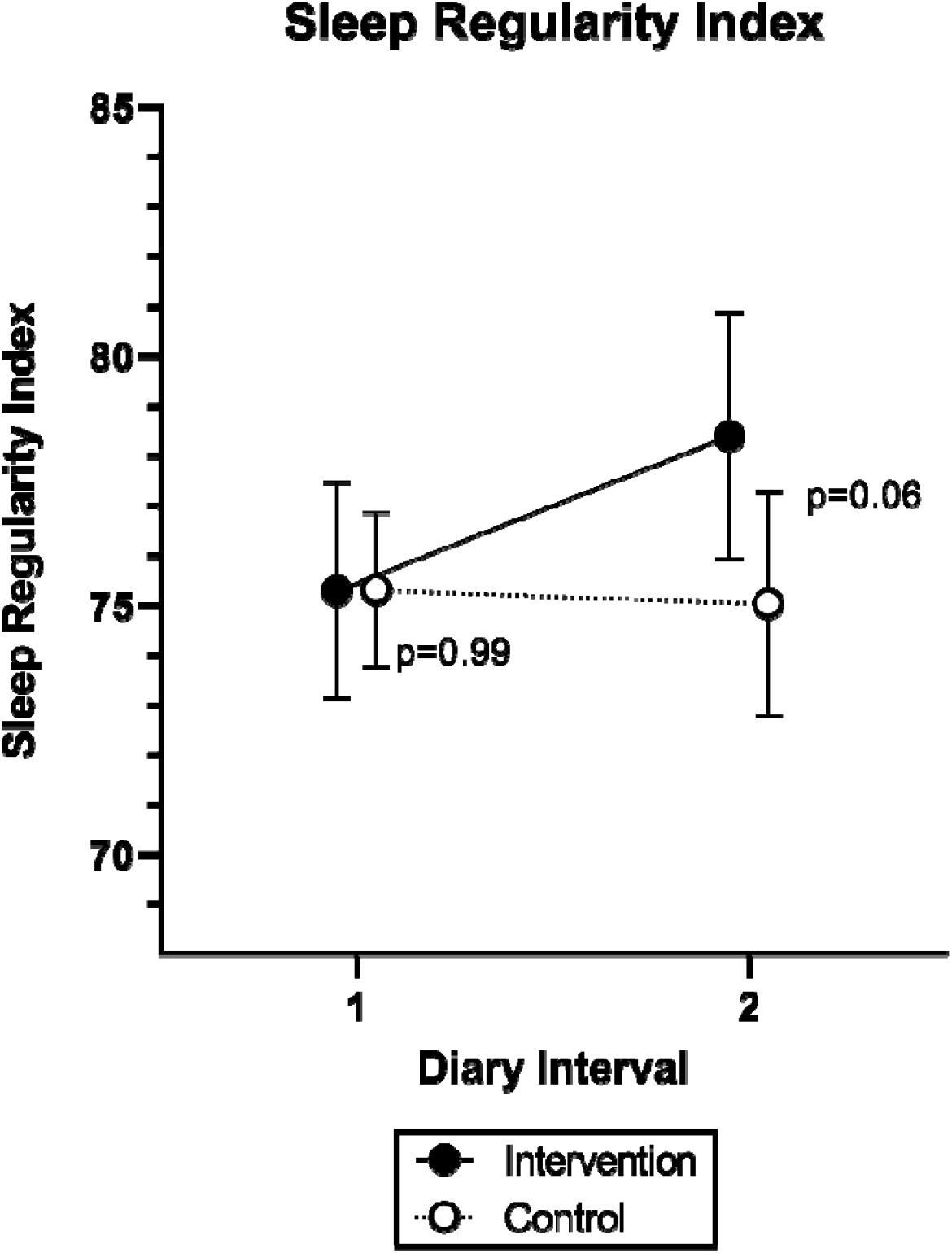
Changes in the Sleep Regularity Index (SRI) during the study between intervention and control conditions.

## Discussion

This randomized clinical trial conducted among employees at a large academic employer in the US Northeast demonstrated a number of favorable sleep, health, and workplace outcomes associated with exposure to sleep health education followed by a personalized smartphone application “dayzz.” Specifically, those randomly assigned to the experimental condition exhibited an increase in healthy sleep behaviors (i.e., less sleepiness, more sleep consistency, and increased sleep duration). We also observed an increase in sleep duration (on both work and free nights) on prospective monthly questionnaires. Those in the intervention condition demonstrated a trend toward a more regular sleep schedule according to the Sleep Regularity Index and a significant increase in overall sleep quality. Regarding workplace outcomes, those in the experimental condition also demonstrated a trend toward less absenteeism and significantly lower presenteeism. Finally, we observed lower healthcare utilization (fewer mental health visits, specifically, and fewer total healthcare visits, broadly) in the experimental condition. In summary, this randomized clinical trial demonstrates that a workplace sleep wellness program can be beneficial to the employee and employer.

## Data Availability

All data produced in the present study are available upon reasonable request to the authors.

